# UShER-TB: Scalable, Comprehensive, Accessible Phylogenomic Analysis of *Mycobacterium tuberculosis*

**DOI:** 10.1101/2025.07.22.25331806

**Authors:** Lily M Karim, Francisco José Martínez-Martínez, Ash O’Farrell, Angie S. Hinrichs, Theo Sanderson, Zamin Iqbal, Varvara K. Kozyreva, John M. Bell, Mariana G. López, Iñaki Comas, Russell Corbett-Detig

**Affiliations:** Department of Biomolecular Engineering, University of California, Santa Cruz; Genomics Institute, University of California, Santa Cruz; Tuberculosis Genomics Unit, Instituto de Biomedicina de Valencia (IBV), CSIC, Spain; Department of Infection Biology, London School of Hygiene and Tropical Medicine, London, UK; Milner Centre for Evolution, University of Bath, UK; Microbial Diseases Laboratory, Infectious Disease Laboratory Division, California Department of Public Health, Richmond, CA; CIBER in Epidemiology and Public Health, Madrid, Spain

**Author notes:** Equal contributions (Lily M Karim, Francisco José Martínez-Martínez).

## Abstract

*Mycobacterium tuberculosis,* the bacterium responsible for the Tuberculosis (TB) disease, remains a leading global infectious disease killer, and genomic epidemiology is essential for understanding its transmission dynamics. Computational limitations prevent comprehensive phylogenetic analysis of the publicly available *Mycobacterium tuberculosis* genomes. Here, we create UShER-TB, a comprehensive pipeline for scalable phylogenomic MTB analysis. We processed 129,312 MTB genomes to construct a comprehensive global phylogeny capturing unprecedented genomic diversity. UShER-TB achieved high accuracy in transmission cluster reconstruction. The comprehensive phylogeny also facilitated identification of putative novel lineages and sublineages, and successful placement of ancient DNA samples. The UShER-TB platform enables real-time phylogenomic analysis of new genomes, revealing transmission hotspots and introduction patterns at global scales. Our approach overcomes longstanding computational barriers, providing researchers with efficient tools for TB genomic surveillance especially for resource-limited settings where TB burden is highest.

## Introduction

Tuberculosis (TB) has been present in human and animal populations for thousands of years. The first solid evidence of the *Mycobacterium tuberculosis* (MTB) pathogen infecting humans dates back at least 8,000 years^1^. Since then, TB has probably killed more people than any other single pathogen^2^. It has been the leading cause of death from a single infectious agent worldwide for decades, only briefly surpassed by COVID-19^3^, and is one of the ten top leading causes of mortality^3,4^. It is estimated that there are 10 million new TB cases and more than 1.2 million deaths annually^3,4^, disproportionately affecting disadvantaged populations. Its burden varies with factors such as overcrowding, HIV status, malnutrition, diabetes and access to healthcare^3^.

Novel genomic tools improve strategies to halt transmission locally and globally. Global strategies implemented in the 1990’s such as directly observed therapy^5,6^ have been successful in reducing deaths from a peak of over 2 million deaths per year. However, the decline in the number of cases has been slow (at a rate of 2% per capita), and the world population growth counteracts this decrease such that overall case numbers remain relatively steady^7^. Current progress is therefore far from the World Health Organization Sustainable Development Goals’ target of ending the TB epidemic by 2030. Transmission remains common even in low-burden countries. An additional issue is the emergence of drug-resistant tuberculosis. While global rates are low (∼3%), some regions report that more than 20% of new cases are multidrug resistant, implying the transmission of already drug resistant strains^8^. Recent reports also show substantial transmission of strains resistant to new drugs recently introduced in treatment programs^9^. Genomic techniques developed in recent years have proven essential to understanding the nature of TB transmission, quantifying TB transmission in different settings, identifying transmission hotspots, enabling outbreak investigations, differentiating between relapse and reinfection, and distinguishing imported versus local cases^10^. In addition, genomic data fosters biological research by allowing us to understand the evolution of the pathogen, its phylogeographic spread, and its co-evolution with human populations^11^.

The large MTB genome size, with 4.4 million base pairs, makes these analyses very time-consuming as the number of samples increases. With current tools, phylogeny inference can take several days to finish with just a few thousand samples. More than 250,000 short-read MTB datasets are publicly available on the Sequence Read Archive (SRA), however. This means that most available data must be omitted from analyses and chosen for inclusion *a priori*, even though this may accidentally exclude relevant data. Although sample subsetting is performed based on scientific evidence (e.g. previous epidemiological studies), this potential bias may obscure new insights into transmission.

During the COVID-19 pandemic, the extraordinary pace and scale of generation of SARS-CoV-2 genome sequences inspired the creation of phylogenetic tools like MAPLE (MAximum Parsimonious Likelihood Estimation^12^) and UShER (Ultrafast Sample placement on Existing tRees^13,14^). The UShER tool accepts a pre-existing phylogeny and places new sequences on it, avoiding the need to reconstruct the whole tree from scratch when new samples are analyzed. UShER was successfully used in regional and local genomic epidemiology studies^15,16^, and is currently the tool used to identify SARS-CoV-2 lineages globally as a part of the Pangolin network^14^.

UShER makes it possible to reconstruct massive phylogenies for SARS-CoV-2 with potential applications to diverse other pathogens. Here, we expand this platform to MTB, whose genome is approximately 150X the size of SARS-CoV-2. Using UShER to study MTB shows it is a valuable tool for genomic epidemiology studies in bacteria: 1) it has potential to use most publicly available data, 2) complex computational analysis is handled in a user-friendly way, and 3) it is fast and efficient, thereby enabling application in resource-limited settings and offering rapid results. In this study we detail (1) the new algorithms developed to adapt UShER for MTB genomic epidemiology, (2) the creation of a comprehensive UShER online phylogenomics platform including 129,312 available MTB samples, and (3) results obtained from UShER for transmission reconstruction, surveillance, analysis of ancient DNA samples, and potential for public health applications. Hereafter, we refer to the complete product, workflows, new methods and resulting datasets as “UShER-TB”. Collectively, this work develops tools and resources to build trees with as many MTB samples as needed in an efficient way and to place new samples in those trees, creating a new paradigm for phylogenetic and transmission studies.

## Results

### The myco bioinformatic pipeline for processing MTBC whole genome sequences

We developed a comprehensive platform for genotyping and phylogenetic analysis of newly sequenced MTB genomes. This resource consists of a bioinformatic pipeline available from Dockstore (https://dockstore.org/workflows/github.com/aofarrel/myco/myco_sra; https://dockstore.org/workflows/github.com/aofarrel/myco/myco_raw) and GitHub (https://github.com/aofarrel/myco) an expanded web platform for visualization and interpretation (https://taxonium.org/tuberculosis/SRA; usher.bio), and a curated publicly accessible and nearly comprehensive phylogenetic tree for MTBC(https://hgdownload.gi.ucsc.edu/hubs/GCF/000/195/955/GCF_000195955.2/UShER_Mtb_SRA/). The pipeline is designed to be modular, and it is possible to run its components separately. This was done to account for technical limitations of the compute platforms used by some public health jurisdictions that necessitate splitting certain parts of the pipeline into multiple components. A full explanation of version differences can be found in Supplementary Materials, but in short, we recommend using TBD_sra for users looking to generate a phylogenetic tree from NCBI SRA samples, or TBD_raw if inputting FQ files directly (https://dockstore.org/workflows/github.com/aofarrel/TB-D/TBD_sra).

### A comprehensive whole-genome MTBC phylogeny

MTBC is a worldwide respiratory pathogen of high public health importance. There are hundreds of thousands of available whole genome sequences. As a consequence of technological limitations, these have not been aggregated into a single comprehensive phylogenetic tree. To meet that need, we considered all paired-end samples categorized as MTBC available on the SRA at the time (June 2024), representing over 200,000 samples. Taking into account both manual identification of problematic samples and automated quality-checking via the myco pipeline, as well as removal of some miscategorized samples and samples incompatible with our pipeline, we generated TBProfiler and variant information for 135,181 samples (SupplementalTable1.tsv) and inferred a phylogenetic tree. We removed 5,732 samples (5,632 from FRS screening (SupplementalTable2.tsv) and 100 from manual inspection (SupplementalTable3.tsv)) that may be mixtures of two or more genetically distinct lineages (see Methods) and pruned 137 *M. canettii* samples (SupplementalTable4.tsv), leaving a total of 129,312 samples on the final phylogenetic tree. The global phylogeny is encoded in a binary file^13^ that is currently 17Mb in size. To test placement efficiency, we randomly selected 100 samples from the dataset, duplicated them and added them to the tree as new samples. With 40 designated threads, this addition took 3 minutes and 11 seconds and used a peak of 7.1 GB of RAM.

### Rich metadata for the global phylogeny

Accurate and rich sample metadata is the most impactful resource for improving the utility and interpretability of genomic datasets and databases^17^. We developed Ranchero, a set of tools and resources for obtaining standardized metadata for diverse samples. Using this tool, we processed all 242,430 BioSamples tagged as belonging to the *Mycobacterium* genus as of September 2024. Ranchero standardized NCBI SRA metadata fields for the entire genus, as well as additional sample metadata from a variety of publications^18,19,20,21,22,23,24,25,26,27,28,29,30,31^ (SupplementalTable5.tsv), which includes information not present in some samples’ NCBI metadata fields, such as sampling locations and dates. This includes experimentally derived phenotypic drug susceptibility testing (pDST) data (SupplementalTable6.tsv) for 31,552 samples. The resulting genus-wide metadata file is provided as SupplementalTable7.tsv. This rich metadata ensures that the UShER MTBC tree is maximally interpretable for a wide variety of applications in public health, biomedical, and evolutionary research aimed at understanding and controlling TB transmission.

From the SRA and publications (see methods) 71,741 (55.48%) of the 129,312 samples placed on the final phylogenetic tree are annotated with a host organism, the most common(n=66,827, 93.15%) being human, while 3,687 (5.14%) were labeled as cattle/bovine, including buffalo and bison. The next most common hosts were badger (n=382, 0.53%), deer (n=306, 0.43%, including elk but excluding antelope)), and pig/boar (n=176, 0.25%), respectively. Additionally, 49.69% (n=64,250) of samples are accompanied by metadata that identifies the year of isolation.

In addition to collecting SRA metadata, Ranchero includes TBProfiler and general QC information for samples processed by our myco pipeline. From this, we determine that our samples typically had a median coverage above 50 (median = 53, mean = 52.37). Out of the 129,312 samples on our phylogenetic tree, TBProfiler assigned lineages to 99.88% (n=129,162) samples. TBProfiler sometimes applies more than one lineage per sample; a total of 128,960 samples were given a single unambigious main lineage by TBProfiler. Out of these unambigious lineages, the most commonly assigned was lineage 4 (n=57,109, 44.28% of data with one lineage), followed by lineage 2 (n=38,864, 30.13%), lineage 1 (n=11,442), and lineage 3 (n=11,300). The most common of the three animal-adapted lineages that TBProfiler can assign was La1 (*M. bovis*, n=8,276) followed by La3 (*M. orygis*, n=268) and La2 (*M. caprae*, n=184). TBProfiler additionally assigned 489 and 967 as the *M. africanum* lineages 5 and 6 respectively. The rarer lineage 7 (n=39), lineage 9 (n=27), and lineage 8 (n=3) were all present as well. This information is also present in the genus-level metadata file (SupplementalTable7.tsv), along with lineages provided in NCBI metadata tags and/or existing publications. TBProfiler additionally provided drug-resistance metadata for all samples (n=129,314) on the tree, which is labeled separately from the subset of data for which we have pDST. The most common drug resistance delineation from TBProfiler was Sensitive at 51.47% (n=66,561), followed by MDR-TB (indicating multiple drug resistance) at 18.25% (n=23,595). Notably, 11.95% (n=15,447) of samples were marked as "Other," more than Pre-XDR-TB (n=11,688), HR-TB (9,255), RR-TB (2,412), and XDR-TB (356) delineations, showing the limitations of commonly-used shorthands when describing drug resistance.

### Availability of the comprehensive MTBC UShER resources

The final tree (Figure 1A) and associated sample metadata (partially summarized in Figure 1B) are available in several formats for visualization and sample placement. First, we developed a Taxonium build to host the tree for visualization and inspection of sample metadata (available from <u>Taxonium</u>^32^). Second, we host the tree for download in various formats as a part of the UCSC Genome Browser (https://hgdownload.gi.ucsc.edu/hubs/GCF/000/195/955/GCF_000195955.2/UShER_Mtb_SRA. Finally, as a part of the UShER.bio project users can place newly genotyped samples on the phylogenetic tree on demand at no cost (https://usher.bio).

**Figure 1:**
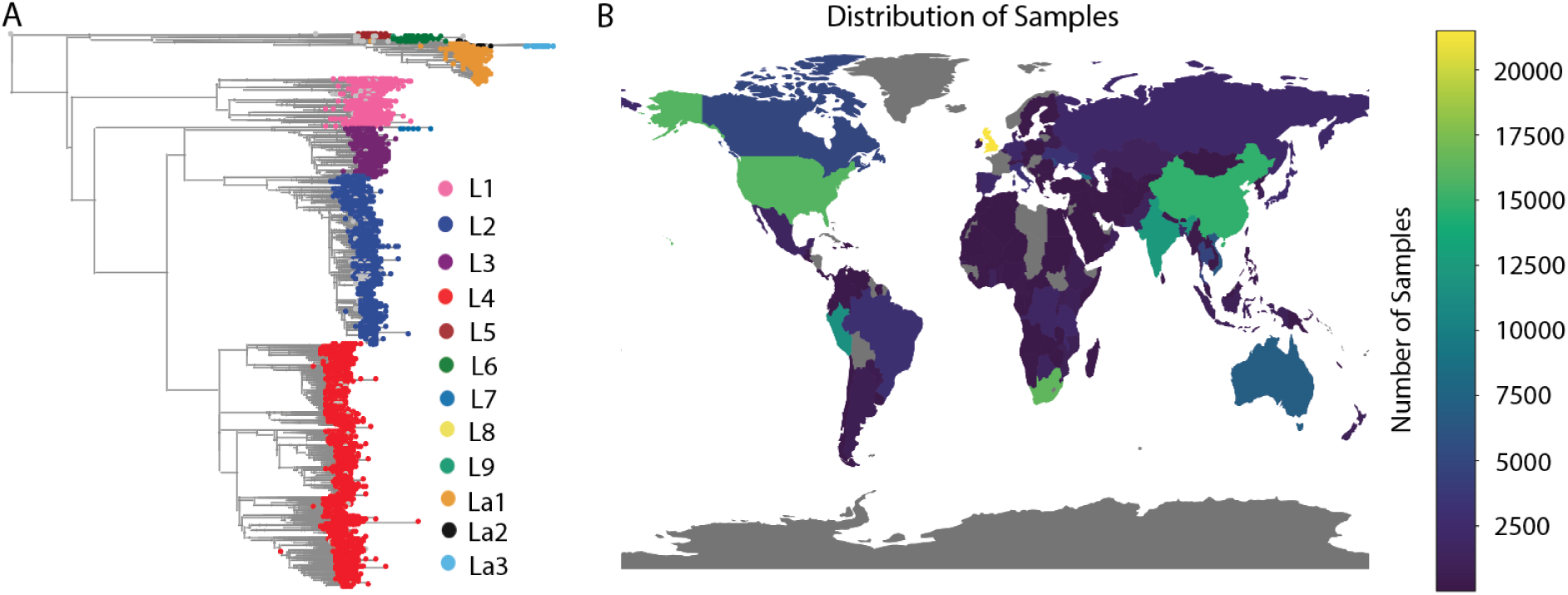
Summary of samples included in the global MTBC UShER phylogenomics resource. A) Global phylogeny of 129,312 samples processed with the myco pipeline and placed with usher-sampled. The phylogeny, samples and metadata can be explored interactively on <u>Taxonium</u>. B) Geographical distribution of samples. Note that sample depth does not necessarily correspond to density of infections, as sequencing resources contribute heavily to data collection.

### Evaluation of phylogenetic inference accuracy and utility

To assess whether UShER’s placement accuracy is sufficient for this MTBC resource to be used for detailed public health investigations into local outbreaks, we benchmarked the approach. One common analysis goal is to identify samples that are closely related to newly sequenced genomes^33–35^. We simulated data with properties similar to the global dataset (see methods) and computed the “neighborhood recovery rate” (hereafter NRR; see methods), a metric that asks what proportion of samples within a given divergence time are discovered using a particular SNP-distance threshold. As expected, a larger SNP threshold can recover larger subsets of closely related samples (ExtendedDataFigure1.svg) and this NRR can be quite sensitive. For example, with a true divergence time of 2 years or fewer, a threshold of one SNP is able to recover all closest relatives in 84.3% of cases (ExtendedDataFigure1.svg). Therefore, a relatively small SNP threshold can accurately identify the closest relatives and will minimize the number of additional samples considered. Nonetheless, when new samples are obtained from geographic regions with little genomic data and correspondingly few close relatives are discovered, a larger threshold may be necessary.

In addition to this fine-scale NRR metric, we find that UShER-sampled is able to accurately estimate the molecular evolutionary history of the species — i.e., the phylogenetic tree. The normalized Robinson-Foulds distance between the true and inferred phylogeny is 0.03 (where 0 would indicate identical topologies, and 1 completely discordant phylogenies). We also calculated RF between a phylogeny inferred by IQ-Tree-2 and UShER-TB for the three real-world datasets enriched in transmission cases used in the later sections. For the Valencia Region the normalized RF distance is 0.04, for Thailand it is 0.01 and for Moldova 0.11. All three values are quite modest, indicating good topological agreement between inference methods. This indicates that phylogenetic relationships are usually reconstructed by UShER-sampled for real MTB genomic datasets. More generally, we conclude that these resources will be a valuable addition to the public health and evolutionary genomics toolkits for investigating MTBC evolution and transmission.

### Post phylogenetic inference processing of locally missing data

Missing data can significantly hinder accurate phylogenetic reconstruction for genomic datasets (see Methods, Figure 2A). In public health settings, where SNP thresholds are often used to identify closely related samples, it’s crucial to exclude positions where a sample lacks a confident genotype. A common approach is to mask alignment positions with missing data in some samples. However, with the large number of samples in our analysis, this would result in masking nearly the entire genome.

**Figure 2:**
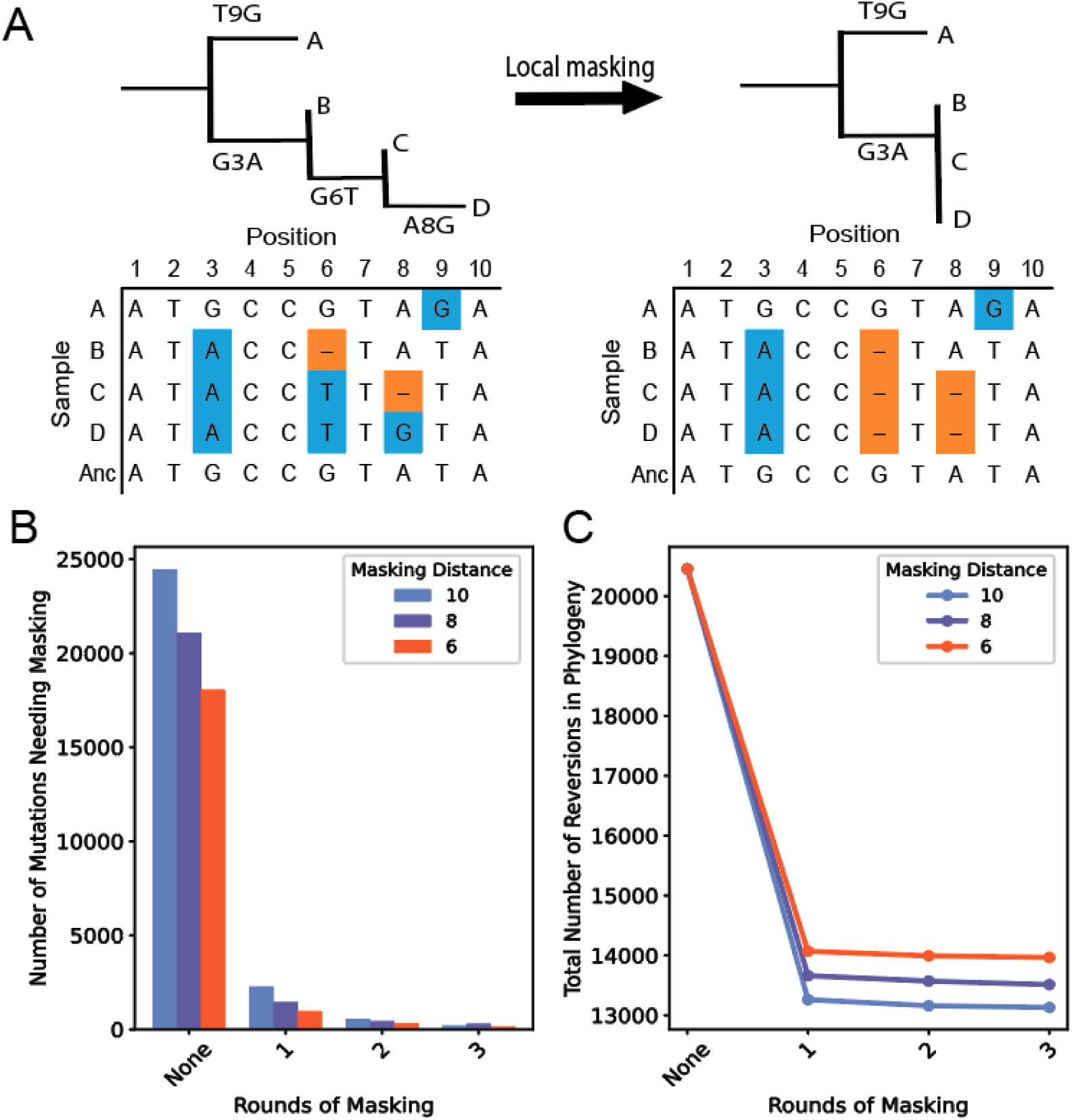
Motivation and effects of local masking. A) Example demonstrating a placement artifact that occurs when samples have missing sequence information, where their close neighbors may have variants. On the left, Sample B has missing data (highlighted in the sequence alignment in orange) at position 6, while neighbors C and D share a >T mutation (mutations highlighted in blue) there. The placement algorithm treats this as a difference, inflating mutation distances despite alignment evidence. To avoid such false distances between closely related samples, we mask mutations like those at positions 6 and 8 (right)(see Methods). B) To test the effect of multiple rounds of post inference local masking, we tracked the number of mutations needing to be masked across 4 rounds of the procedure. C) We tracked the number of revertant mutations present in the global tree after each round of masking to assess improvements in phylogenetic quality.

To address this, we developed a method to only mask locally missing data. Rather than masking sites across all samples, we only mask positions in a given sample if a closely related sample on the tree also lacks a confident genotype (see Methods, Figure 2A). Because masking can alter the tree and introduce new missing data patterns, we applied multiple rounds of this local masking after phylogenetic inference to see if further improvements could be made. In each round, we measured the number of mutations that needed to be masked and the number of revertant mutations (mutations that mutate back to the ancestral allele)^36^. Although revertant mutations can occur naturally, the occurrence of reversions within a genomic dataset are more likely due to assembly or sequencing error (a sign of inaccurate phylogenetic reconstruction) ^36^.

After just one round of local masking, the number of mutations to be masked dropped by 91–94%, depending on the SNP distance threshold used (between 6 and 10). Revertant mutations also dropped by 41–45% under the same conditions. Using a SNP distance of 10, we masked 24,436 SNPs across 15,946 nodes, and found that a single round was sufficient to identify the majority of problematic sites (Figure 2B and C).

Notably, 11.2% of the masked sites contained revertant mutations—compared to just 0.63% of all mutations across the tree—suggesting that our approach effectively removes inaccurately inferred sites (Figure 2C).

### Transmission cluster assignment in local and global datasets

We benchmarked usher-sampled against a standard pipeline for identifying transmission clusters based on pairwise distances using a 10-SNP threshold. Considering both approaches, we obtained a unique list of 769 samples in transmission at a maximum of 10 SNPs, of which 705 (91.7%) are shared in both methods. Considering the number of transmission clusters, we obtained a unique list of 230 clusters, 176/230 (76.5%) that match between both methods. Out of the 54 non-matching clusters, 35/230 (15.2%) only show 1 or 2 samples of difference between both methods, most discrepancies can be corrected by simple visual inspection of the phylogenetic topology and branch lengths. Finally, 19/230 (8.3%) clusters were only identified by the standard pipeline. Cluster files from the standard pipeline and obtained after using usher-sampled are available at https://github.com/lilymaryam/Mtb_global_phylogeny.

Having established that usher-sampled is accurate to identify most transmission clusters, we further explored usher-sampled’s utility for reconstructing genomic transmission clusters using real data from different settings and in different analysis scenarios, tailored by public health needs. Clustering is challenging as it depends on low SNP thresholds (typically from 0 to 10)^37^, and thus any change in the analysis pipeline may affect the results. In our case, we estimated transmission clusters based on phylogenetic distances (see Methods) after *de novo* building reference phylogenies for each dataset.

As diverse transmission rates in the different settings may influence transmission cluster reconstruction, we estimated the proportion of samples in transmission for each dataset using a 10 SNPs threshold. In the Valencian Region, Chiang Rai province and Moldova, transmission measured as genomic clustering rates accounted for 46.5% (n=716/1540, 193 clusters), 37.1% (n=433/1167, 119 clusters) and 62.3% (n=1210/1925, 245 clusters), respectively.

Considering this variety of settings, we evaluated the accuracy of placement for usher-sampled. At a local scale, i.e. mimicking ongoing regional surveillance, usher-sampled placed new samples with high accuracy and showed strong agreement with the original clusters. When using a global phylogenetic framework, the tool maintained cluster integrity while combining local and global context, further validating its robustness. Finally, in a global phylogeny with no previous genomic context, which simulates a real-world scenario of recently-established genomic surveillance programs, usher-sampled showed a high concordance with the original clusters. Overall, across all regions and situations, the tool reliably positioned new samples near their original locations and preserved transmission group assignments (Figure 3A). Cluster assignment for all samples in every case can be checked in SupplementalTable8.tsv.

**Figure 3:**
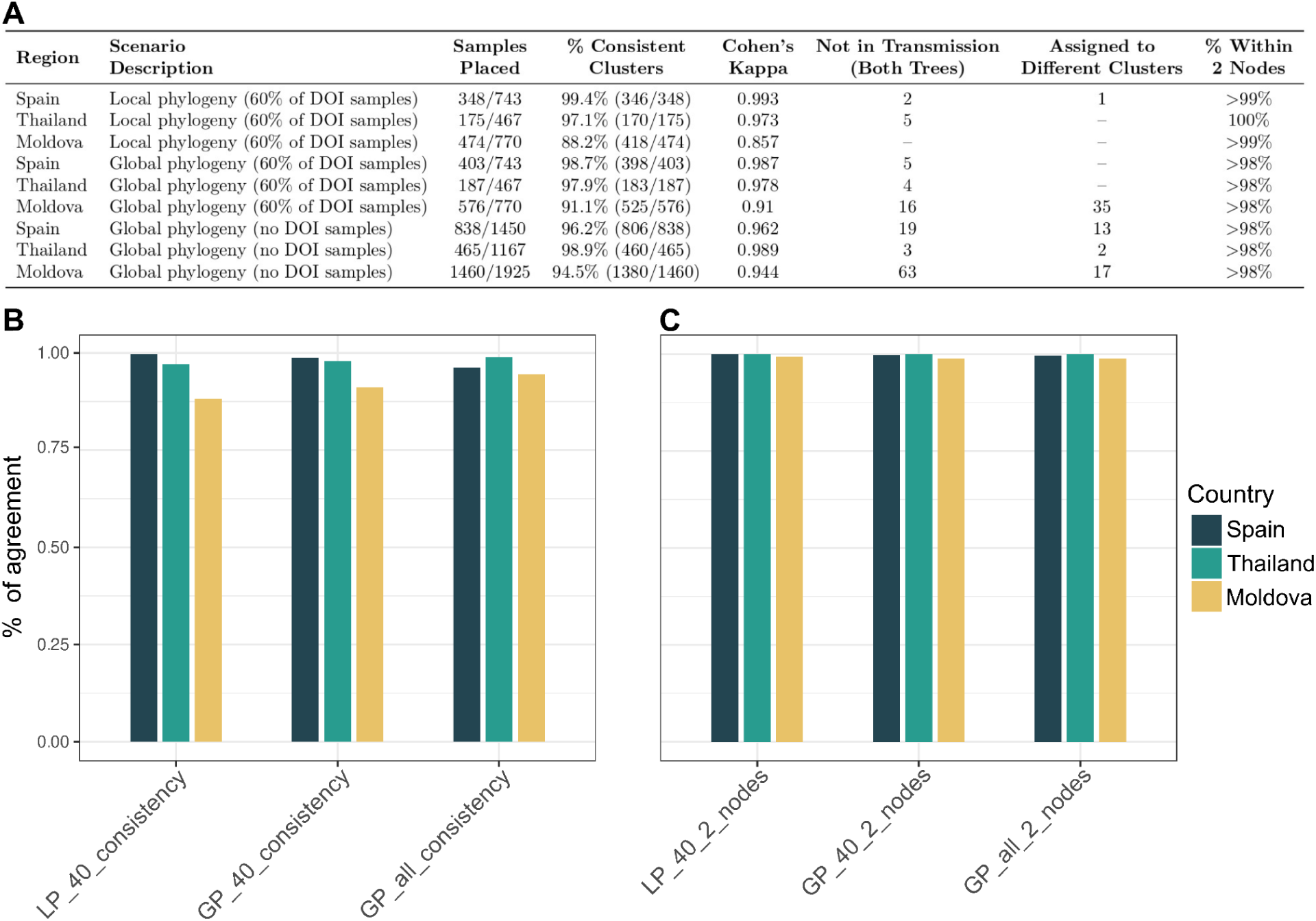
Placement consistency in different settings. A) Summary of transmission cluster assignment results in local and global settings. B) Proportion of samples that are correctly assigned to their original transmission group. C) Proportion of samples that fall within 2 or less nodes from their original location in the tree. In both cases, different scenarios are considered. From left to right: local dataset placing the 40% of samples (LP_40), global dataset placing the 40% of the samples (GP_40) and global dataset placing all the sequences (GP_all).

### Demonstration of applicability for global epidemiological spread

A key strength of comprehensive phylogenomic resources such as this is the capacity to illuminate dynamics of global pathogen spread. In particular, one important component is the frequency of introduction of pathogen lineages into new geographic regions — which necessarily requires genomic sequences accompanied by geographic metadata as we have assembled. We investigated the utility of our resources for this purpose by calling new lineage introductions using a pandemic-era tool^38,39^. We identified 51,077 total introductions, which we subsequently categorized as “singletons” (where only one sample supports an introduction event into a given region) and “multiples” (where two or more samples in a given region are closely related and likely to descend from a single introduction, potentially indicating ongoing transmission after introductions) (ExtendedDataFigure2.svg). This analysis revealed distinct introduction patterns among lineages, which are presumably related to the geographic endemic regions of each lineage and the geographic origin of the migrant flows related to each country but are also likely influenced by biases in sequencing efforts. This result highlights the utility of UShER-TB for identifying global transmission dynamics and will serve as a critical resource for such investigations in the future.

### Identification of new candidate lineages

The scale of the global phylogeny is such that it may allow for the discovery and characterization of new lineages and sublineages. By creating a comprehensive global phylogeny, we can carefully inspect samples across the tree that have not been successfully assigned lineage metadata (Figure 4). Among these unlabeled samples, we have found several that carry unique genetic variation not present in more widespread lineages. Sample SAMEA7423516 is sister to Lineage 5 with a branch length (467) comparable to the branch lengths of the samples within Lineage 5 (Two-Sided MWU, p∼0.15 (see Methods), and passed all of our quality control filters, including those intended to eliminate mixed infection samples. Sample SAMEA3448377 appears among the most genetically divergent samples in Lineage 3, potentially representing a previously unknown sublineage. This Malawian sample has 353 unique mutations that distinguish it from its most recent common ancestor with other samples within Lineage 3. We found no significant difference (p∼0.09, Mann-Whitney U Test) between the branch length of SAMEA3448377 and sister samples in Lineage 3, showing that SAMEA3448377 was not basally placed in Lineage 3 and therefore is unlikely to be a mixed infection. Furthermore, the sample passed our quality control filter for mixed bases.

**Figure 4:**
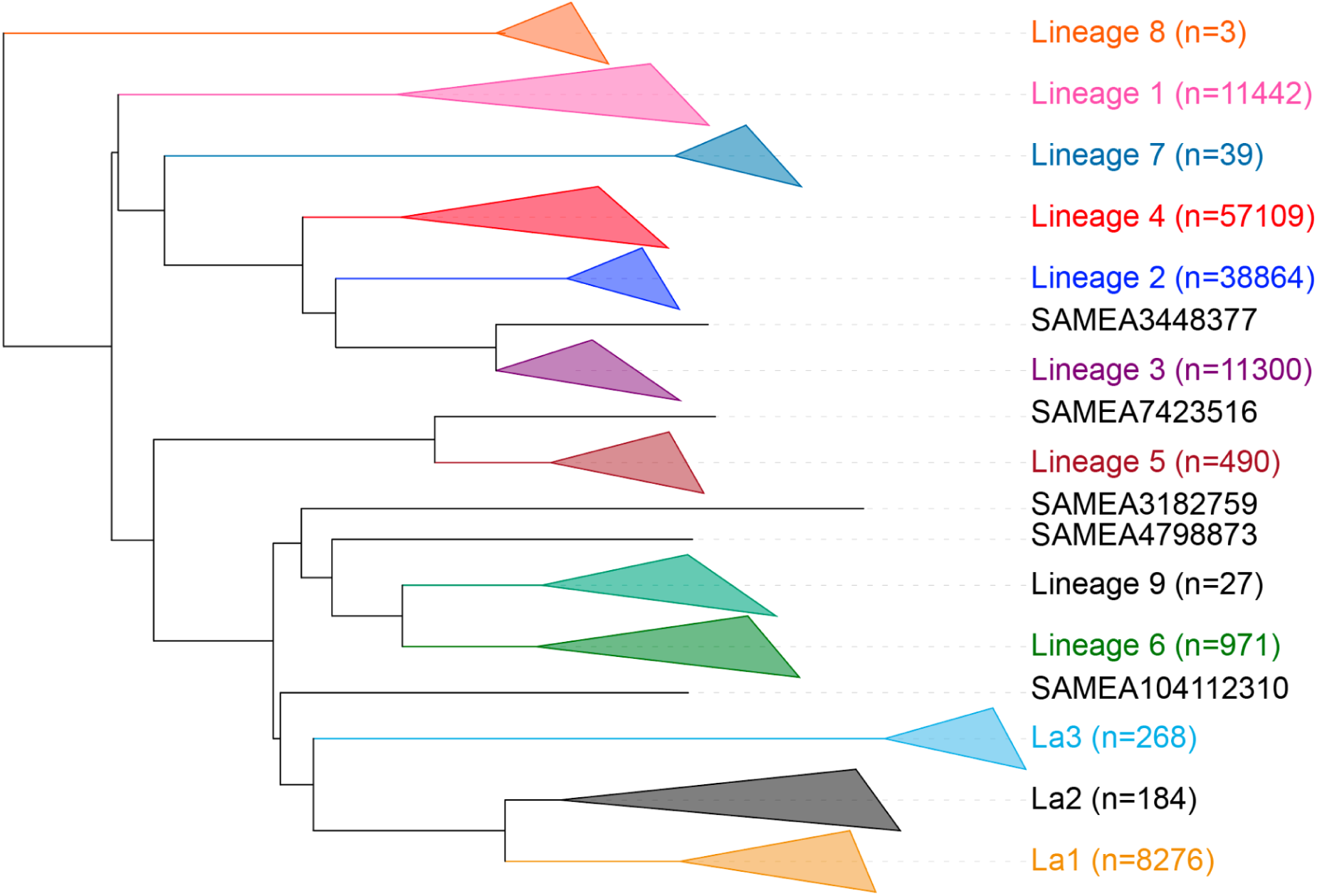
Location of new candidate lineages. Summary of the UShER-TB global tree (with major lineages collapsed), illustrating where samples with potential novel lineage designation are phylogenetically placed.

Several samples, including SAMEA3182759 and SAMEA4798873, are paraphyletic outgroups to lineages 6 and 9. These samples likely belong to a previously characterized animal-adapted MTBC clade A1^19^. SAMEA104112310 is part of a small clade that is sister to the animal-hosted lineages La1-La3. This group of samples, including SAMEA104112310, appears to be part of an animal-hosted clade A2 that includes infections characterized as *M. microti* (land mammal hosted) and *M. pinnipedii* (pinniped hosted)^19^. These and other samples outside of major lineages demonstrate the utility of our global phylogenetic resource for comprehensively documenting the vast genetic variation across MTBC.

### Phylogenetic placement of ancient DNA pathogen samples

Current techniques have paved the way to sequence DNA from strains infecting individuals that lived hundreds of years ago. This allows us to better calibrate the evolution of the bacteria and the disease, improving our knowledge of how it emerged, how it evolved and its epidemiology in the past. UShER-TB is capable of assigning ancient DNA samples to their correct background. For example, MTB samples from mummies sampled in Hungary (SAMEA2810262 and SAMEA2810266) are placed in nodes enriched in L4.1.2.1 and L4.8/L4.10, respectively (Figure 5A and B), matching their original lineage typing^40^. An MTB genome obtained from a 17th Century Swedish Bishop^41^ (SAMN14570812) is also assigned to the correct L4.8/L4.10 clade(Figure 5C). As for genomes from Peruvian pre-contact human skeletal remains, they are thought to be close^42^ to an animal clade related to *M. pinnipeddii*, associated with seals and sea lions. UShER-TB, however, places the samples in a basal position to the common clade between *M. pinnipedii* and *M. microti* sequences, instead of closer to *M. pinnipeddii* (Figure 5D). After reconstructing a maximum-likelihood tree with IQ-TREE containing the strains of the monophyletic node in which the new samples are placed, we retrieved the expected topology for this region of the phylogeny as published in Bos et al.^42^ (ExtendedDataFigure3.svg). The example illustrates that analysis of most ancient DNA samples is technically challenging and will need a tailored pipeline, which is beyond the scope of this study. Nonetheless, our results show that UShER-TB can perform initial screening of the samples, genotyping and identify their phylogeographic background for detailed downstream analyses.

**Figure 5:**
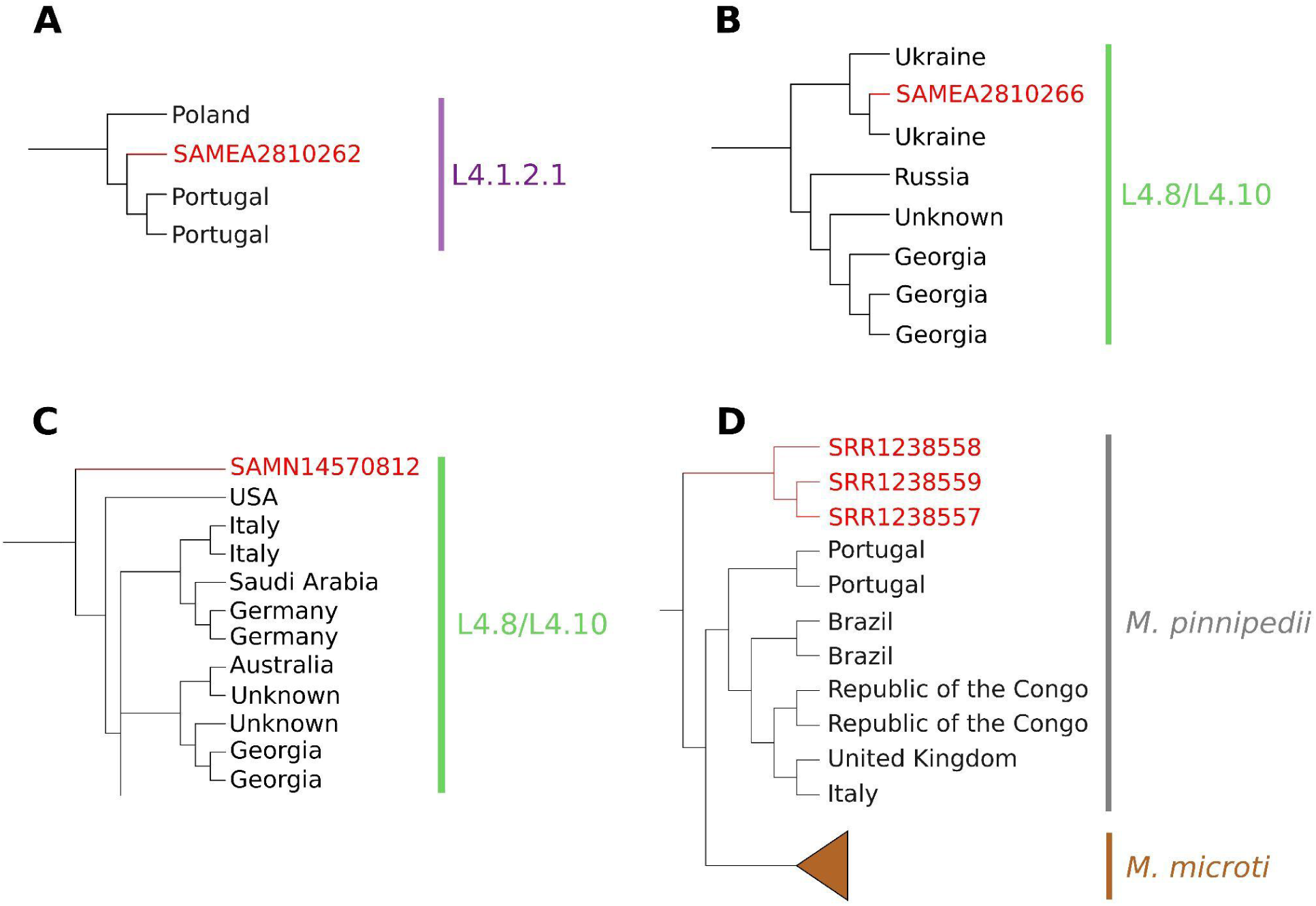
Phylogenetic neighbourhoods of ancient DNA-placed samples. All the nodes show the placed samples (in red), the neighbouring taxa codified by the country the samples were collected in, and the lineage of the samples within the node. A) and B) correspond to the Hungarian mummies’ sequences, C) corresponds to the Bishop’s sequence and D) shows the *M. pinnipedii* sequences.

## Discussion

This study establishes a globally comprehensive genomic resource for *Mycobacterium tuberculosis*, addressing long-standing challenges in TB phylogenetics and transmission inference. By leveraging large-scale publicly available sequencing data and implementing efficient tree-building methodologies, this work enables the analysis of MTB genomes at an unprecedented scale. The ability to rapidly place new genomes into a global phylogeny provides a more nuanced understanding of TB transmission dynamics, helping to distinguish between recent and historical transmission events with greater confidence. These improvements have practical implications for public health, particularly in settings where genomic surveillance has been constrained by computational limitations or data fragmentation.

Nonetheless, several important caveats must be considered when interpreting results from UShER-TB. First, we recommend that new samples be processed using uniform bioinformatics pipelines, as differences in variant calling, quality filtering, or read mapping approaches may introduce artifacts that affect phylogenetic placement accuracy and comparability. Second, sampling remains highly non-uniform globally, with particularly sparse representation from high-burden regions in Africa and other low-resource settings, which can complicate epidemiological interpretations when few phylogenetically close relatives are available for newly sequenced isolates, a limitation common to phylogeographic studies. Third, our current approach focuses exclusively on single nucleotide polymorphisms and does not capture structural genomic variation, including large insertions, deletions, or rearrangements as well as regions inaccessible to short-read mapping that may significantly impact MTB phenotypes such as antibiotic resistance^43–45^. Future integration with pangenome approaches (such as Walia et al.^46^) and an increasing use of long-read technologies may help address this limitation by incorporating structural variation into comprehensive phylogenomic analyses^47^. These considerations underscore the importance of continued efforts to expand global genomic surveillance and to refine analytical methodologies for MTB.

Beyond its immediate epidemiological applications, this resource also enhances broader research efforts into TB evolution, drug resistance emergence, and phylogeographic patterns. By integrating a vast number of genomes into a single analytical framework, this approach minimizes biases introduced by selective subsampling and allows for more representative insights into MTB population structure. Moreover, its computational efficiency ensures that researchers in a wide range of settings, including those with limited resources, can access and utilize these tools effectively^48^. As TB control strategies increasingly incorporate genomic data, the methods and dataset presented here provide a foundation for more comprehensive and timely investigations into the spread and adaptation of this persistent pathogen.

## Online Methods

### Data acquisition

We considered almost all publicly-available Illumina-processed MTBC (*Mycobacterium tuberculosis* Complex) sequences on the SRA^49^ as of June 2024, which we queried using the using the SRA Cloud Based Metadata Table (https://www.ncbi.nlm.nih.gov/sra/docs/sra-cloud-based-metadata-table/).

We curated metadata from fields associated with each sample’s SRA entry, or in some cases, by parsing supplemental material of publications. In particular, we obtained sample metadata through direct parsing of NCBI fields, pulled using BigQuery (https://www.ncbi.nlm.nih.gov/sra/docs/sra-bigquery-examples/), existing publications (SupplementalTable5.tsv), and expert consultation (E. Roycroft and J. Andrews pers comm.). We note that there is a wide variety of metadata fields on SRA, which are not always consistent. Other issues can arise when comparing across samples that use different formats for date-of-isolation, or older taxonomic/lineage systems. To aid in this, as well as to harmonize metadata parsed from individual publications, we developed an auxiliary tool called Ranchero (github.com/aofarrel/ranchero). Although designed for wrangling tuberculosis data, most of Ranchero’s functions are compatible with other organisms.

### Sample processing

We passed sample accessions to our MTBC processing pipeline, myco (https://dockstore.org/workflows/github.com/aofarrel/myco/myco_sra), which extracts sequence data from the SRA as paired-end fastq files and generates variant information. The myco pipeline uses SRANWRP (https://github.com/aofarrel/sranwrp), a wrapper for sra-tools^50^ we developed to streamline the mass-download of BioSamples that include at least one valid Illumina paired-end sample, while also automating the exclusion of entries with incompatible data formats or corrupted fastq files. The myco pipeline uses clockwork^51,52^, a CRyPTIC consortium^18^ data processing pipeline, for accurate genotyping of short-read sequencing of tuberculosis samples^52–56^. We downsampled FASTQ files above 450 megabytes to 1,000,000 reads using <u>seqtk sample</u>^57^, and discarded samples with fewer than 1000 reads. We masked regions of the genome that are known to be difficult to genotype^58^ using the CRyPTIC mask file^18,59^. Our pipeline converts low-depth regions and indel/structural variants to ambiguous nucleotides so that neither are used for placement by the UShER algorithm, and performs several QC checks (see details in https://github.com/lilymaryam/Mtb_global_phylogeny). For each QC passing SRA BioSample, version 6.2.4 of myco generated a clockwork^52^-processed bed coverage file and VCF^60^ (Variant Call Format), the latter of which is converted into MAPLE file format^12^ using the genome H37Rv (NC_000962.3) as the reference. The MAPLE file format^12^ is used for downstream analyses because VCF is inefficient for genomic epidemiological applications at this scale^12^. Our pipeline produces TBProfiler^61,62^ (version 4.4.2) reports, which provide lineage designations and predicted drug resistance, for each sample. These reports are parsed and used as metadata for the final tree, along with the other metadata standardized by Ranchero. (SupplementalTable7.tsv, see Results).

### Phylogenetic inference

Using 135,290 samples processed with the myco pipeline as input, we ran UShER (version 0.6.4) subtool, usher-sampled^14^, to infer and optimize a global MTB phylogenetic tree with default parameters. We used TaxoniumTools^32^ (PyPi version 2.0.86) for visualizing the initial tree.

### Identification of potential coinfections

After inferring the initial global phylogeny, we identified and removed samples putatively coinfected with two or more genetically distinct strains. We used the VCFs generated from myco to obtain adjudicated variant calls and Filtered Read Support (FRS) information^52^. FRS indicates the percentage of reads that support a given variant call. Because ambiguous nucleotides and missing data are preferentially resolved to reference by usher-sampled when more than one equally parsimonious assignment is possible^13^, coinfected samples are expected to place near to the common ancestor of coinfecting lineages. We identified all samples where greater than 19% (>0.19) of its variants were classified as having a low FRS (<0.85) and labeled these samples as ‘candidate coinfections’ (SupplementalTable3.tsv). We selected this threshold by examining the visual representation of the data, optimizing it to efficiently prune low-quality samples near ancestral nodes without discarding high-quality data. (ExtendedDataFigure4.svg). We then manually pruned 100 additional samples due to their phylogenetic placements near to deep internal nodes on the phylogeny, which is not a plausible placement for modern samples (SupplementalTable4.tsv).

### Rerooting and pruning *M. canettii*

To reroot our global phylogeny, we first identified the *M. canettii* clade^63^. Though *M. canettii* is a member of the MTBC, it has significantly different biological properties and is often used as an outgroup^19,64,6519,20,66^. We rooted the tree at the common ancestor of Lineage 8 and *M. canettii*. We inserted a node corresponding to the reconstructed ancestor in the final tree and pruned the 137 *M. canettii* samples (SupplementalTable5.tsv).

### Post inference local masking

Missing sequence information in the genomic dataset can cause issues during phylogenetic reconstruction. Missing data can obscure true genetic similarity between samples, especially when neighboring samples have mutations at those sites. This can lead placement algorithms to overestimate their differences. To mitigate this, we mask such positions to reduce the impact of misleading signals. We describe these issues in Figure 2A, with an example phylogeny. We developed an approach that reduces the impact of missing data on phylogenetic reconstruction (see Figure 2A) that we call post-placement local masking. Our method searches the tree for sample pairs within a nominal “masking distance”, which is their phylogenetic distance in SNPs. It then identifies all missing positions in a given sample and masks those sites in all other samples within the masking distance. By looking only within other closely related samples, we are able to mask locally. We then use matOptimize ^67^ to rearrange the tree to optimal parsimony after masking. Our masking tool is available in the matUtils library^38^. This method may improve the analysis of transmission between closely related samples because it does not arbitrarily resolve genotypes at missing sites.

### Evaluation of the accuracy of UShER for public health inquiry

We simulated a 30,000 sample MTB genomic dataset, constructed a phylogeny, and evaluated UShER’s accuracy. To do this, we used Alisim^68^ to simulate a large genomic dataset of MTB genomic samples. We extracted a subtree of 30,000 neighboring samples from the inferred global phylogeny (see above) and input this subtree as the guide tree in Alisim (hereafter the “truth tree”), the length of the H37Rv (NC_000962.3) genome as alignment length, the General Time Reversible (GTR)^69^ model with MTB base frequencies (A:0.18, C:0.32, G:0.32, T:0.18) as our evolutionary model. We emphasize that although the tree based on real data must contain inference errors, once we fix a topology as truth for simulation of genomic data, those errors are no longer relevant. After we reconstructed the phylogeny using usher-sampled^14^, we calculated the normalized Robinson-Foulds distance^70^ between the truth tree and the inferred tree after collapsing internal nodes that do not contain mutations (as in Kramer et al.^71^). This approach is appropriate because some nodes are multifurcating and would be arbitrarily resolved in binary trees, which is typical in densely sampled genomic epidemiology applications^71,72,73^.

We also evaluated the accuracy of sample placements in relation to their closest neighbors as a typical public health analysis goal is to find all known samples with a recent shared common ancestor. We devised a test that computes the percentage of the truth neighbors that can be recovered within a specified distance of the target sample in the inferred tree. We extracted neighboring samples from each tested sample in the truth tree based on the expected maximum age of divergence of neighboring samples (i.e. a 2 year divergence neighborhood is expected to contain samples within 2 SNP of our target^42,74–77^). For the analysis, we extracted truth neighborhoods with divergence ages of 1 year, 3 years, 5 years, 8 years, and 10 years. In the inferred tree, we extracted SNP neighborhoods of distances 1, 5, and 10 from around the target sample because they are standard transmission cluster sizes that epidemiologists use for analysis^10,75,76,77,21^. For each divergent neighborhood, we calculated the proportion of true nearest neighbors in the inferred tree, which we call the Neighborhood Recovery Rate or NRR (ExtendedDataFigure1.svg).

### Estimating the number of introductions

To find novel introductions into specific geographic regions, we used matUtils^39^. We used matUtils’ annotate function to add lineage metadata to the global phylogeny (SupplementalTable9.tsv). We then ran matUtils introduce on the annotated phylogeny, which identifies all putative geographical introductions, with default settings and a metadata file associating each sample with its associated ISO code (SupplementalTable10.tsv). We separated the introduction count per country into multiple transmissions and single introductions, and stratified these categories by lineage (ExtendedDataFigure2.svg).

### Identification of potential new lineages

We identified potential new lineages through visual inspection of the 129,312 sample tree. For each lineage, we searched for unannotated samples that were placed basally. To rule out the possibility that these samples’ unusual phylogenetic placements are due to a mixed infection, we calculated the path lengths from internal node to tip for every sample descending from the most recent common ancestor (MRCA) of these samples and samples in the sister clade (in both cases, the parent of the sample was the MRCA). We compared these distributions with a 2-sided Mann-Whitney U test to determine if the branch lengths are significantly different between all descendant samples. Furthermore, we note that these samples were not excluded based on mixed-base criteria, suggesting that their unusual phylogenetic positions are not due to coinfection or contamination.

### Transmission cluster assignment in local and global datasets

Tuberculosis transmission surveillance is key for public health systems to evaluate transmission burden and to investigate outbreaks. In this context, we assessed the capacity of usher-sampled to reconstruct known transmission clusters, defined as groups of MTB samples with a pairwise genetic distance of a maximum of 10 SNPs. We chose this because it is a common SNP threshold in MTBC cluster analysis^78^.

To benchmark the robustness between a standard, widely used pipeline for cluster identification (available at https://gitlab.com/tbgenomicsunit/ThePipeline) based on pairwise distances and assignment using usher-sampled, we used a dataset from the Valencian Region in Spain (n=1540). We used the standard approach for cluster identification using a 10 SNPs threshold and compared them to usher-sampled clusters using a R script (available at https://github.com/lilymaryam/Mtb_global_phylogeny), the non-matching clusters were visually inspected to identify potential sources of discrepancy between both approaches.

After validating the use of usher-sampled to identify transmission groups, we evaluated the approach in two different scenarios. The first scenario is whether sequences from a local setting will be analyzed alone as part of ongoing surveillance programmes in a country. The second scenario is when those sequences are incorporated into a large global reference phylogeny. In both cases, we evaluated the accuracy of phylogenetic placement using usher-sampled and an existing input phylogeny. Three settings with different sampling scheme, burden and transmission patterns were chosen: the aforementioned Valencian Region in Spain, a low-burden setting (<10 cases per 100.000 population, collected in the period 2014-2019, n=1540), Chiang Rai province in Thailand (157 cases per 100.000 population in 2023, collected in the period 1999-2011, n=1167) and Moldova (74 cases per 100.000 population, collected between January 2018 and December 2019, n=1925).

To generate a ground truth for each of the settings, we built a local and a global phylogeny (SupplementalTable11.tsv) using usher-sampled for de novo phylogenies. Then, using a custom script in R (https://github.com/lilymaryam/Mtb_global_phylogeny) we obtained the pairwise genetic distance between all samples and used this matrix as input for another custom Python script (https://github.com/lilymaryam/Mtb_global_phylogeny) that obtains the clusters for a given SNP threshold, 10 in our case, which was the basis for our comparison. To test whether sample placement can assign samples consistent with known transmission clusters, we removed some of the samples from each of the datasets. In the Valencian Region (n=1540), collected between 2014 and 2019, we removed the samples collected after 2016 (n=743). In Thailand (n=1167), collected between 1999 and 2011, and Moldova (n=1925), collected between 2008 and 2011, we removed 40% of the samples (n=467 and n=770, respectively) by sorting all samples by collection date and excluding the most recently collected ones. A scheme on how each dataset is compared to the ground truth can be found in Figure 6.

**Figure 6:**
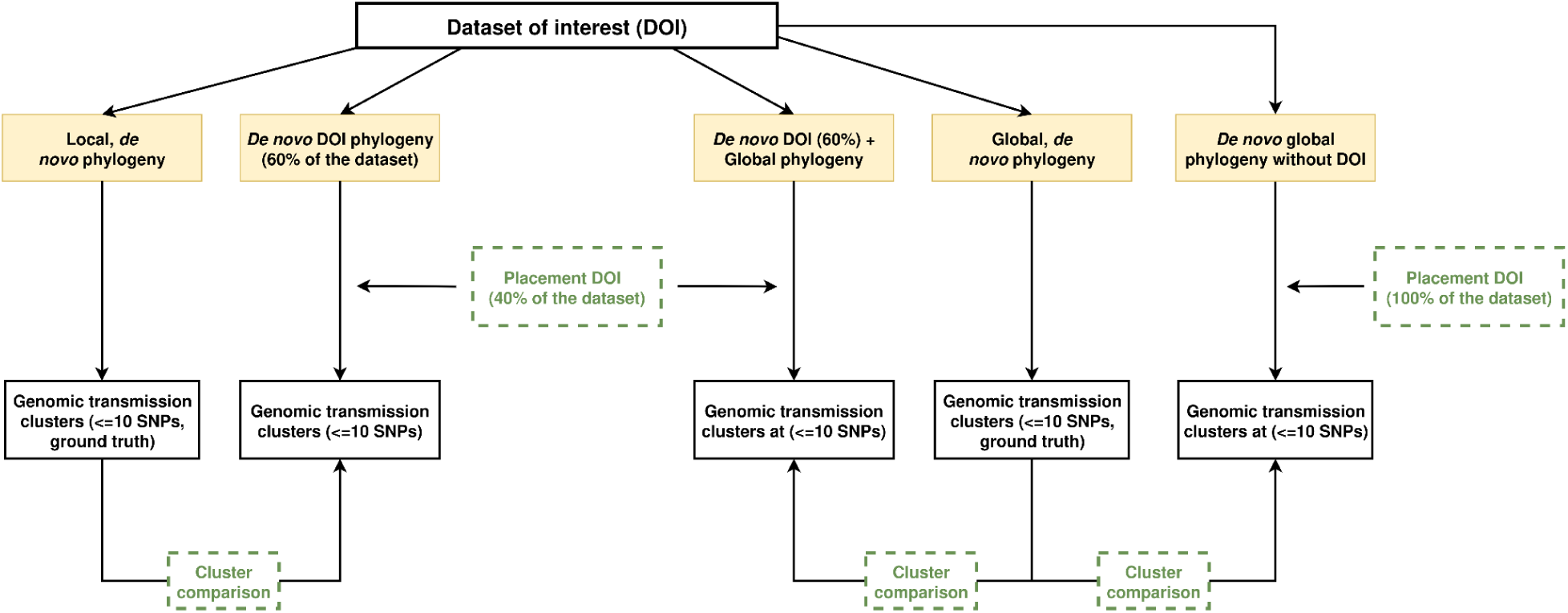
Transmission cluster assignment comparison workflow.

### Ancient MTBC genomes placement with usher-sampled

We used usher-sampled to place six ancient DNA genomes from three different studies and timeframes. Among 14 genome sequences from Hungarian mummies dating back to the eighteenth century, we chose those two with the highest coverage and no signature of mixed infection^40^. We also included a recently sequenced MTB genome obtained from skeletal remains of a Swedish bishop born in the seventeenth century^41^. Finally, we included three 1,000-year-old sequences, obtained from Peruvian human skeletons, closely related to extant *Mycobacterium pinnipedii* strains^42^. In the first 2 ancient datasets (Hungarian, Swedish), the myco pipeline was used as described above. However, the Peruvian dataset did not meet the predefined myco quality standards, i.e., having less than 20% of positions with a coverage below 10, and thus was discarded by the pipeline. As a result, a different variant-calling pipeline was used to analyze these sequences. In short, the pipeline calls the variants using the VarScan and Mutect2 callers^79,80^, and performs a variant consensus with Minos^52^. The final variants we use for placement have a depth of at least 20 reads and a minimum base quality of 15. Then, the output is converted to VCF using the faToVcf tool from the UShER toolbox and finally to MAPLE format using a custom script (https://github.com/lilymaryam/Mtb_global_phylogeny) so it can be placed with usher-sampled. After the placement of the six ancient samples, we used IQ-TREE to further reconstruct a local phylogeny of the monophyletic clade where these samples were placed to evaluate whether the placement performed by usher-sampled in a global context was able to retrieve the expected phylogenetic relationships seen at a local scale, also for ancient DNA samples.

## Code availability

We provide major workflow components to support our conclusions and enable future applications here. Specifically, two versions of the myco pipeline are available -- myco_raw, which takes in fastqs as direct file inputs, and myco_sra, which takes in a text file of BioSample accessions from which fastqs are downloaded. myco’s components are designed to be modular (such as SRANWRP, which pulls fastqs from SRA) and may be useful for analysis of other organisms. We additionally provide Tree Nine, a workflow which takes in the MAPLE-formatted diff files output by either flavor of myco and places them on a phylogenetic tree.

myco_sra and myco_raw share a GitHub repository at https://github.com/aofarrel/myco, and have Dockstories entries at https://dockstore.org/workflows/github.com/aofarrel/myco/myco_sra and https://dockstore.org/workflows/github.com/aofarrel/myco/myco_raw respectively. Tree Nine can be found on Dockstore at https://dockstore.org/workflows/github.com/aofarrel/tree_nine/tree_nine or GitHub at https://github.com/aofarrel/tree_nine/tree_nine.

DOI versions of these workflows are also available:

- myco_sra: https://doi.org/10.5281/zenodo.15080116
- myco_raw: https://doi.org/10.5281/zenodo.15061623
- Tree Nine: https://doi.org/10.5281/zenodo.15046888

Custom scripts for analysis and visualizations presented in this work are available via GitHub from: https://github.com/lilymaryam/Mtb_global_phylogeny.

## Data availability

All genotypic data used in this work are available from the SRA. See SupplementalTable7.tsv for detailed information about specific BioSamples.

As described in our Methods, metadata -- including pDST, pipeline outputs, and sample collection information -- was sourced from a variety of open sources. We used ranchero to standardize and compile all gathered metadata in TSV format, available in SupplementalTable6.tsv Note that this file is derived from a very wide BigQuery search of SRA, as described in SupplementalFile1.txt, and includes many samples not present on the tree. A version of our metadata subset only to samples on the tree can be found in SupplementalFile12.tsv. The tree, annotated with a subset of our metadata, may be viewed using Taxonium: https://taxonium.org/tuberculosis/SRA. The tree may be downloaded from our supplemental drive https://drive.google.com/drive/folders/1Tzzmh3Y1WXe_cLmdHZIToppyGvfvJqNL or from https://hgdownload.gi.ucsc.edu/hubs/GCF/000/195/955/GCF_000195955.2/UShER_Mtb_SRA/ All supplemental files are available at https://drive.google.com/drive/folders/1Tzzmh3Y1WXe_cLmdHZIToppyGvfvJqNL?usp=sharing

## Funding Acknowledgements

This work has been supported by the following: Centers for Disease Control 75D30124C20302, and California Department of Public Health NU50CK000539 to RC-D; European Research Council (ERC): H2020-ERC-COG/0800, Ministerio Español de Ciencia e Innovación: PID2022-137607OB-I00 to I.C.; individual fellowship PRE2020-094308 to F.M; Wellcome Trust 306920/Z/23/Z to T.S.; NHGRI Predoctoral Fellowship T32HG012344 to L.K.

## Data Availability

As described in our Methods, metadata -- including pDST, pipeline outputs, and sample collection information -- was sourced from a variety of open sources. We used ranchero to standardize and compile all gathered metadata in TSV format, available in SupplementalTable6.tsv Note that this file is derived from a very wide BigQuery search of SRA, as described in SupplementalFile1.txt, and includes many samples not present on the tree. A version of our metadata subset only to samples on the tree can be found in SupplementalFile12.tsv. The tree, annotated with a subset of our metadata, may be viewed using Taxonium: https://taxonium.org/tuberculosis/SRA. The tree may be downloaded from our supplemental drive https://drive.google.com/drive/folders/1Tzzmh3Y1WXe_cLmdHZIToppyGvfvJqNL or from https://hgdownload.gi.ucsc.edu/hubs/GCF/000/195/955/GCF_000195955.2/UShER_Mtb_SRA/ https://drive.google.com/drive/folders/1Tzzmh3Y1WXe_cLmdHZIToppyGvfvJqNL https://github.com/lilymaryam/Mtb_global_phylogeny https://hgdownload.gi.ucsc.edu/hubs/GCF/000/195/955/GCF_000195955.2/UShER_Mtb_SRA/ https://taxonium.org/tuberculosis/SRA

